# Are psychiatric disorders risk factors for COVID-19 susceptibility and severity? a two-sample, bidirectional, univariable and multivariable Mendelian Randomization study

**DOI:** 10.1101/2020.11.29.20240481

**Authors:** Jurjen J. Luykx, Bochao D. Lin

## Abstract

Observational studies have suggested bidirectional associations between psychiatric disorders and COVID-19 phenotypes, but results of such studies are inconsistent. Mendelian Randomization (MR) may overcome limitations of observational studies, e.g. unmeasured confounding and uncertainties about cause and effect. We aimed to elucidate associations between neuropsychiatric disorders and COVID-19 susceptibility and severity. To that end, we applied a two-sample, bidirectional, univariable and multivariable MR design to genetic data from genome-wide association studies (GWASs) of neuropsychiatric disorders and COVID-19 phenotypes (released on 20 Oct. 2020). In single-variable Generalized Summary MR analysis the most significant and only Bonferroni-corrected significant result was found for genetic liability to BIP-SCZ (a combined GWAS of bipolar disorder and schizophrenia as cases vs. controls) increasing risk of COVID-19 (OR = 1.17, 95% CI, 1.06-1.28). However, we found a significant, positive genetic correlation between BIP-SCZ and COVID-19 of 0.295 and could not confirm causal or horizontally pleiotropic effects using another method. No genetic liabilities to COVID-19 phenotypes increased risk of (neuro)psychiatric disorders. In multivariable MR using both neuropsychiatric and a range of other phenotypes, only genetic instruments of BMI remained causally associated with COVID-19. All sensitivity analyses confirmed the results. In conclusion, while genetic liability to bipolar disorder and schizophrenia combined slightly increased COVID-19 susceptibility in one univariable analysis, other MR and multivariable analyses could only confirm genetic underpinnings of BMI to be causally implicated in COVID-19 susceptibility. Thus, using MR we found no consistent proof of genetic liabilities to (neuro)psychiatric disorders contributing to COVID-19 liability or vice versa, which is in line with at least two observational studies. Previously reported positive associations between psychiatric disorders and COVID-19 by others may have resulted from statistical models incompletely capturing BMI as a continuous covariate.

## Introduction

Several large population-based studies have investigated associations between positive testing for COVID-19 on the one hand and psychiatric disorders on the other ^1-3^. Positive test result likelihoods for psychiatric disorders are inconsistent between those studies: while two of those cohort studies (from the UK and South Korea) do not report positive associations between COVID-19 testing and psychiatric disorders ^1,4^, others (from the UK and the US) mention odds ratios of 1.5 to 10 for associations between mental disorders and a COVID-19 diagnosis ^2,3,5^. One observational study reports bidirectional associations between psychiatric diagnosis in the previous year and COVID-19 ^2^. For a recent diagnosis of a mental disorder in the US^2^, odds ratios for COVID-19 were reported to be around 7.6, with evidence for relatively severe COVID-19 outcomes in those with a diagnosis of mental disorder^5^. One limitation of these studies is that psychiatric diagnoses were grouped by some ^2,4^, e.g. as mental disorders, psychotic disorders or affective disorders, precluding conclusions about COVID-19 risks for specific psychiatric disorders. Additionally, some diagnoses, such as bipolar disorder, were not included in some of the analyses ^3^. Furthermore, correction for medical comorbidities decreased several high odd ratios^5^. Finally, some authors admit residual socioeconomic factors may not be sufficiently captured in some databases^2^. Indeed, as recently noted, confounding or biases (partly) explain associations of COVID-19 with a range of traits and diseases ^6^. Mendelian Randomization (MR) has the potential to overcome two major limitations of observational studies: unmeasured confounding and uncertainties about cause and effect. Examples of MR studies elucidating risk factors for COVID-19 include two recent studies showing that BMI and smoking are risk factors for COVID-19 ^7,8^. We are not aware of preprinted or published MR studies of psychiatric disorders and COVID-19. The recently updated whole-genome data on COVID-19 phenotypes (https://www.covid19hg.org/results/, see methods section) further increases the timeliness of MR approaches to elucidate risk factors for COVID-19 diagnosis and severity.

We hypothesized that given the aforementioned inconsistent observational evidence and attenuation of reported odds ratios when including covariates, psychiatric disorders do not constitute strong risk factors to contract COVID-19 or develop a severe course of COVID-19. Similarly, we hypothesized that genetic liability to COVID-19 would not increase susceptibility to psychiatric disorders. To test our hypotheses, we performed a range of bidirectional univariable and multivariable MR analyses of genetic liability to major psychiatric disorders and to COVID-19 susceptibility as well as severe COVID-19.

## Methods

### Overview

We performed 2-sample mendelian randomization (MR) using summary statistics from large genome-wide association study (GWASs) to examine whether genetic liabilities to (neuro)psychiatric disorders increase risk of contracting COVID19 and of a severe course of COVID-19 (forward MR analyses, considering (neuro)psychiatric disorders as exposures and COVID-19 as outcome). In addition, we used COVID-19 GWASs to examine potential effects of genetic liabilities to COVID-19 diagnosis and to COVID-19 severity on (neuro)psychiatric diseases (reverse MR analyses, considering COVID-19 as the exposure and (neuro)psychiatric phenotypes as the outcomes). The principle of MR analyses is shown and further explained in **Supplementary Figure 1**. Most of our methods outlined below have been previously explained in more detail ^9^. The GWAS summary statistics we used were drawn from studies that had obtained written informed consent from participants and received ethical approval from institutional review boards. No ethical approval for the current analyses was needed as they were based on publicly available summary statistics.

### (Neuro)psychiatric summary statistics

We used the available GWASs with summary statistics for psychiatric disorders including Alzheimer’s dementia (AD) (a meta-analysis of the stage 1 discovery dataset ^10^), anxiety ^11,12^, anxiety and stress-related diagnoses (ASRD) ^11,12^, Major Depressive Disorder (MDD) ^13^, Bipolar disorder (BIP) ^14^, Schizophrenia (SCZ) ^15^, and a meta-analysis of BIP and SCZ (BIP-SCZ; **Table 1**)^16^. To avoid that sample overlap between exposures’ and outcomes’ datasets impacted results substantially by inducing instrument bias in MR analyses, we excluded UK Biobank (UKBB) cohorts from (neuro)psychiatric disorders summary statistics. The largest Anorexia Nervosa GWAS and the largest 2018 and 2019 MDD GWASs contained a large number of UKBB participants, precluding us from using those GWASs for MR analyses. Hence, we selected MDD summary statistics from the 23andme cohort^13^, containing only 10,000 independent SNPs due to participants consent, and thus used these summary statistics only for univariable forward MR analysis. Similarly, we refrained from using the psychiatric cross-disorder GWAS as its study population was also partly composed of UKBB participants. For anxiety and ASRD, we performed meta-analysis in METAL ^17^ excluding UKBB participants: of anxiety using the iPSYCH (4,584 cases and 19,225 controls) and ANGST cohorts (7,016 cases and 14,745 controls) and of ASRD also using the iPSYCH cohort (4,584 cases and 19,225 controls) and ANGST cohorts (12,665 cases and 14,745 controls). We used the anxiety and ASRD summary statistics we had thus generated for our MR analyses. ^18^

**Table 1.** GWASs of (neuro)psychiatric disorders used for the current study and outcomes used for the univariable forward MR analyses. In bold are depicted the (neuro)psychiatric GWASs that had identified ≥2 genome-wide significant loci and were thus selected as exposures in our forward MR analyses.

| GWASs | Cohorts | Number of loci | Number of cases | Number of controls | Outcomes used for univariable forward MR analyses (see table 2 for definitions of A-D) |
| --- | --- | --- | --- | --- | --- |
| Anxiety | iPSYCH+ ANGST | 0 | 11,600 | 33,970 |  |
| ASRD | iPSYCH + ANGST | 1 | 19,681 | 33,970 |  |
| **AD** | **ADGC, CHARGE, ADI, ERAD/PERADES.** | **32** | **21,982** | **41,944** | **A1, A2, B1, B2, C1, C2, D1** |
| **MDD** | **23andme (10k SNPs)** | **5** | **75,607** | **231,747** | **A1, A2, B1, B2, C1, C2, D1** |
| **BIP** | **PGC** | **6** | **53,555** | **54,065** | **A1, A2, B1, B2, C1, C2, D1** |
| **SCZ** | **PGC+CLOZ-UK** | **138** | **40,675** | **64,643** | **A1, A2, B1, B2, C1, C2, D1** |
| **BIP-SCZ** | **PGC+CLOZ-UK** | **96** | **20,129** | **21,524** | **A1, A2, B1, B2, C1, C2, D1** |

### COVID-19 summary statistics

We used the most recent COVID19 (GWAS) meta-analyses round 4 (A1, C1, D1; Table 2) and 5 (A2, B, B2, C2, Table 2; round 5 being an updated release from 18 January 2021, with more cohorts and larger sample sizes compared to round 4 only for phenotypes A2, B1, B2 and C2) ^19^, released on October 20, 2020, and January 18, 2021, respectively, from the COVID-19 Host Genetics Initiative (https://www.covid19hg.org/results/) containing several COVID-19 phenotypes. This GWAS was based on a study population drawn from multiple cohorts, with European being the dominant ancestry: BioMe, FinnGen, Genes & Health, LifeLines Global Screening Array, LifeLines CytoSNP, Netherlands Twin Register, Partners Healthcare Biobank, and UKBB (**Table 2; Supplementary Table 1)**. The majority of the included subjects were of European descent, with a small proportion of other ethnic backgrounds. Only variants with imputation quality > 0.6 had been retained and meta-analysis of individual studies had been performed with inverse variance weighting by the authors of the COVID-19 Host Genetics Initiative. To avoid that sample overlap between exposures’ and outcomes’ datasets impacted results substantially by inducing instrument bias in MR analyses, we only included COVID-19 summary statistics that had excluded the 23andme cohort. We divided the COVID-19 phenotypes (A-D, **Table 2**) into two categories, COVID-19 susceptibility and severity. We used two phenotypes to index COVID-19 susceptibility, namely C (defined by the COVID-19 Host Genetics Initiative as partial susceptibility) and D (self-reported COVID-19). Similarly, we used A (very severe respiratory confirmed COVID-19, here: ‘very severe COVID-19’) and B (hospitalized lab confirmed COVID-19, here ‘severe COVID-19’) to index COVID-19 severity.

**Table 2.** Phenotype definition of the COVID-19 phenotypes by the authors of the COVID-19 Host Genetics Initiative used for the current study and outcomes used for the univariable reverse MR analyses. The numbers listed are for the GWASs conducted excluding the 23andme cohorts that were used for the current analyses. For further detail please see https://www.covid19hg.org/results/. In bold are listed the COVID-19 GWASs that had identified ≥2 genome-wide significant loci and were thus selected to be used as exposures in our reverse MR analyses.

| GWAS name | Round | Case definition and number of cases | Control definition and number of controls | N loci | Outcomes used for univariable reverse MR analyses |
| --- | --- | --- | --- | --- | --- |
| A1 | 4 | Laboratory-confirmed SARS-CoV-2 infection AND hospitalized COVID-19 AND (death OR respiratory support), N = 269 | Laboratory-confirmed SARS-CoV-2 infection AND not hospitalized for COVID-19 within 21 days after the test, N = 688 | 0 |  |
| **A2** | **5** | **Laboratory-confirmed SARS-CoV-2 infection AND hospitalized COVID-19 AND (death OR respiratory support), N = 5,582** | **Population, N = 709,010** | **8** | AD, ASRD, Anxiety, BIP, SCZ, and BIP-SCZ |
| B1 | 5 | Laboratory-confirmed SARS-CoV-2 infection AND hospitalized COVID-19, N = 3,961 | Laboratory-confirmed SARS-CoV-2 infection AND not hospitalized for COVID-19 within 21 days after the test, N = 10,538 | 0 |  |
| **B2** | **5** | **Laboratory-confirmed SARS-CoV-2 infection AND hospitalized COVID-19, N = 12,888** | **Population, N = 1,295,966** | **7** | AD, ASRD, Anxiety, BIP, SCZ, and BIP-SCZ |
| C1 | 4 | Laboratory-confirmed SARS-CoV-2 infection OR coding/physician-confirmed COVID-19 OR self-reported COVID-19 via questionnaire, N = 11,591 | (Laboratory tested for SARS-CoV-2 infection AND all tests negative) OR self-reported tested negative for SARS-CoV-2, N = 119,904 | 0 |  |
| **C2** | **5** | **Laboratory-confirmed SARS-CoV-2 infection OR coding/physician-confirmed COVID-19 OR self-reported COVID-19 via questionnaire, N = 36,590** | **Population, N = 1,668,938** | **3** | AD, BIP, ASRD, and SCZ |
| D1 | 4 | Self-reported COVID-19, N = 3,204 | Samples with the minimum possible value from the predictive model AND not self-reported COVID-19 positive, N = 35,728 | 0 |  |

N loci = number of genome-wide significant loci; AD = Alzheimer's disease; ASRD = anxiety and stress-related disorders; Anxiety = Anxiety disorders; BIP = bipolar disorder; SCZ = schizophrenia; BIP-SCZ = a combined GWAS of bipolar disorder and schizophrenia vs. controls.

### MR analyses

Data were analyzed between November, 2020, and January, 2021. As our main analysis, we performed univariable MR using GSMR (Generalized Summary Mendelian Randomization) ^20^ implemented in the Genome Complex Trait Analysis (GCTA) software ^21^. For our exposures, we first selected all relevant single-nucleotide variants (SNVs) identified in each GWASs as having reached a selection P-value threshold < 5 × 10^−8^ and being non-duplicate and uncorrelated (10 000 kilobase pairs apart and Linkage Disequilibrium (LD) R^2^ ≤.001). Instrument outliers were identified using HEIDI-outlier test (p <0.01), with the minimum number of instruments required for the GSMR analysis (nsnps_thresh) = 2. In harmonizing exposure and outcome data we removed palindromic SNVs with intermediate allele frequencies, and SNVs with minor-allele frequency (MAF) differences > 0.2 between exposure GWASs and outcome GWASs. We estimated the F-statistic from first-stage regression to evaluate instrument strength, which is defined as the ratio of the mean square of the model to the mean square of the error. The rule of thumb is that a threshold of F < 10 indicates weak instrument strength^22^.

To interpret our results, we advise readers to take into account effect sizes and not focus on p-values. As a cut-off for statistical significance, we Bonferroni corrected two-sided p<0.05 for the number of tests performed in all analyses, i.e. univariable forward (n=35;), univariable reverse MR (n=16), and multivariable forward and reverse (see results section).

Then, Bonferroni-corrected significant results from GSMR were validated by applying as sensitivity analyses several alternative MR models, namely fixed-effect inverse variance– weighted (IVW), MR-Egger ^23^, weighted median-based regression methods ^24^, and MR-PRESSO ^25^ that depend on different assumptions. The harmonized input data from GSMR were used to perform by “TwoSampleMR” R packages ^26^. For significant results, we used the MR-Egger intercept test, Cochran Q heterogeneity test, and MR pleiotropy residual sum and outlier (MR-PRESSO) test to evaluate potential inverse variance (IV) violations, and Causal Analysis Using Summary Effect estimates (CAUSE) ^27^ to estimate horizontal pleiotropy. We also performed leave-one-out analyses to examine whether any high-impact instruments possibly influenced MR results disproportionally.

At last, we conducted multivariable MR(MVMR) analyses ^28^ using the MendelianRandomization R package to examine which phenotypes remained risk factors taking into account pleiotropic effects among exposures. MVRM estimates the effects of each exposure on an outcome adjusting for genetic associations between multiple phenotypes and the exposure. Scenarios one may think of are when a researcher hypothesizes that exposures are related to one another or when one exposure may mediate the relationship between the exposure of interest and an outcome^29^. MVMR does so by using genetic instruments (derived from either summary-level or individual-level data) associated with each of those multiple phenotypes. MVMR is an extension of MR that may prove more useful and reliable in scenarios where three or more exposures may be at play. Examples for the field of psychiatry include MVMR analyses for self-harm^30^ and schizophrenia^9^. We thus considered MVMR a useful method to follow up and corroborate our initial findings. For MVMR analyses, we constructed instruments using SNVs in each of the GWASs meeting our single-variable MR selection criteria, as described above. We used the MVMR extension of the inverse-variance weighted MR method and MR-Egger method to correct for measured and unmeasured pleiotropy. In forward MVMR, we used as exposures SCZ, BIP and AD, while A1, A2, B1, B2, C1, C2, and D1 COVID-19 phenotypes (**Table 1**) were the outcomes. We excluded the following (neuro)psychiatric diseases as exposures in forward MVMR: BIP-SCZ ^16^ as it was highly correlated with SCZ and BIP (with most of the SNVs overlapping with either SCZ or BIP); MDD ^13^ as it did not have enough SNV information (summary statistics containing only 10,000 independent SNPs due to participants consent); anxiety and ASRD ^11,12^ as they did not have enough instrument variables, i.e. <2. In reverse MVMR analyses, we used the three COVID-19 A2, B2 and C2 phenotypes (**Table 2**) as exposures since they had the required number of ≥2 genome-wide significant loci, while outcomes were ASRD, AD, anxiety, BIP, SCZ, and BIP-SCZ. We removed duplicate and correlated SNVs (within 10 000 kilobase pairs; LD R^2^ ≥0.001), resulting in 8 SNPs as COVID-19 instruments and 119 SNPs as (neuro)psychiatric instruments. Statistical significance for MVMR was Bonferroni-corrected for the number of outcomes (see results section). In the end, genetic heritability and genetic correlations between the neuro(psychiatric) disorders, BMI and COVID-19 phenotypes were estimated using LD Score Regression (LDSC) ^31^.

### Sensitivity analyses

To corroborate the robustness and consistency of our findings, we performed two sensitivity analyses. First, we used a more lenient threshold of (P < 1 × 10^−7^) for genetic instruments as a few SNPs passed conventional genome-wide significant level (P < 5 × 10^−8^), only explaining a small amount of the variance in the complex trait, which in turn may increase chances of type-2 error in MR. This method of relaxing the statistical threshold for genetic instruments has been used in psychiatric MR research when few significant SNPs were available ^32-34^.

Second, we performed MV-MR with more exposures since conditions such as obesity, diabetes and heart diseases may increase the severity of COVID-19 and these and others (e.g. educational attainment and cognitive performance) may be associated with (neuro)psychiatric phenotypes. To that end, we ran univariable MR analyses for COVID-19 outcomes as discussed above using the largest GWAS summary statistics for Body mass index (BMI) ^35^, type 2 diabetes (T2D) ^18^, Coronary artery disease (CAD) ^36^, Educational attainment (EA) and Cognitive performance (CP) ^37^, and Cross-disorder psychiatric disorders (CDG, a combined GWAS of 8 psychiatric phenotypes; here CDG can be used as for COVID-19 datasets UKB and 23andme sections had been excluded; **Supplementary Table 2**) ^38^. Then the abovementioned conditions along with the phenotypes mentioned in Table 2 were entered into multivariable MR analysis as exposures. To avoid sample overlap influencing our results, we conducted MR using COVID-19 round 5 summary statistics which excluded both UK Biobank and 23andme cohorts; namely A2, B1, B2, C2. For (neuro)psychiatric disorders, we replaced the summary statistics containing UK biobank participants with MDD^39^ and ASRD^40^ GWASs that exclude 23andme or UK Biobank cohorts.

### Data and code availability

We have made our code publicly available on Github (https://github.com/Bochao1/MR_PSY_COVID19). The datasets we accessed to perform our analyses may be found in the publications that listed as references for the datasets used.

## Results

### Overview

Five neuropsychiatric disorders (numbers of instruments: AD = 32, BIP = 6, MDD =5, SCZ = 137 and BIP-SCZ = 96; total study populations: AD = 63,926; BIP = 198,882; MDD = 307,354; SCZ = 105,318; and BIP-SCZ = 107,620) had enough (≥2) genome-wide loci to perform forward MR analyses (**Table 1**) on the seven COVID-19 phenotypes defined in **Table 2**. Conversely, three COVID-19 phenotypes (numbers of instruments: A2 = 8, B2 = 7 and C2 = 3; total study populations: A2 = 1,308,275, B2 = 969,689, C2 = 1,388,510; **Table 2**) had enough genome-wide loci to perform reverse MR analyses. As the instrument strength was strong (F-statistic in forward and reverse MR analyses ranging from 36.2 to 69.5), we did not find evidence of weak instrument bias.

### Forward MR results of (neuro)psychiatric disorders as potential risk factors for COVID-19 phenotypes

We performed 35 univariable MR tests to examine the potential effects on COVID-19 of genetic liabilities to 5 psychiatric phenotypes with at least 2 significant loci identified. We thus Bonferroni corrected the significance threshold to 0.05/35 = 0.0014. Single-variable GSMR analysis showed that the top MR result was BIP-SCZ (combined GWAS of bipolar disorder and schizophrenia as cases vs. controls): the effect estimate was consistent with increased COVID-19 susceptibility (D1; N=96 instruments; OR, 1.165, 95% CI, 1.062-1.277; P = 0.0012; **Figure 1**). All four sensitivity MR analyses confirmed the direction of effect detected by GSMR (**Table 3**). IVW, weighted median and MR-PRESSO also showed similar p-values to GSMR (**Table 3**). Although MR Egger was not significant (p= 0.247), overall horizontal pleiotropic effects were absent (for intercept of MR Egger: b= 0.015, p = 0.293). The I^2^_GX_ statistics (I^2^_GX_ =1.07%) of MR Egger were substantially smaller than 90%, suggesting substantial bias in the causal estimates due to uncertainty in the genetic associations, resulting in MR-Egger results not being reliable. The Cochran’s Q test in the fixed-effect IVW model (Q statistic = 9.395, p = 0.152) and MR Egger model (Q statistic = 7.278, p = 0.2) suggested that there was absence of heterogeneity in the instrumental variables, which may be the result of true causality rather than violation of instrument variable assumptions. Furthermore, the leave-one-out analysis (**Figure 2A**) showed that no SNVs altered the pooled IVW beta coefficient, confirming the stability of our results. We also found absence of directional pleiotropy and the instrument strength independent of direct effect (InSIDE) assumption to be satisfied (**Figure 2B**)^23^. As we found a genetic correlation between BPSCZ and D1 (rg = 0.26, SE=0.12, p=0.012), horizontal pleiotropy may influence causality effects. Using CAUSE to estimate causal effects and potentially horizontal pleiotropic effects, no significant causal effects (γ = 0.04, 95% CI=-0.03-0.11) or horizontally pleiotropic effects (η = 0.1 95% CI=-0.79 −1.07) were identified. When comparing the sharing and causal models, the Δ ELPD > 0 (Δ ELPD = 0.022; SE = 0.835; P = 0.51), indicating the posteriors from the causal model predict the data better, so the causal model may be a better fit (Supplementary **Figure 2**).

**Figure 1.**
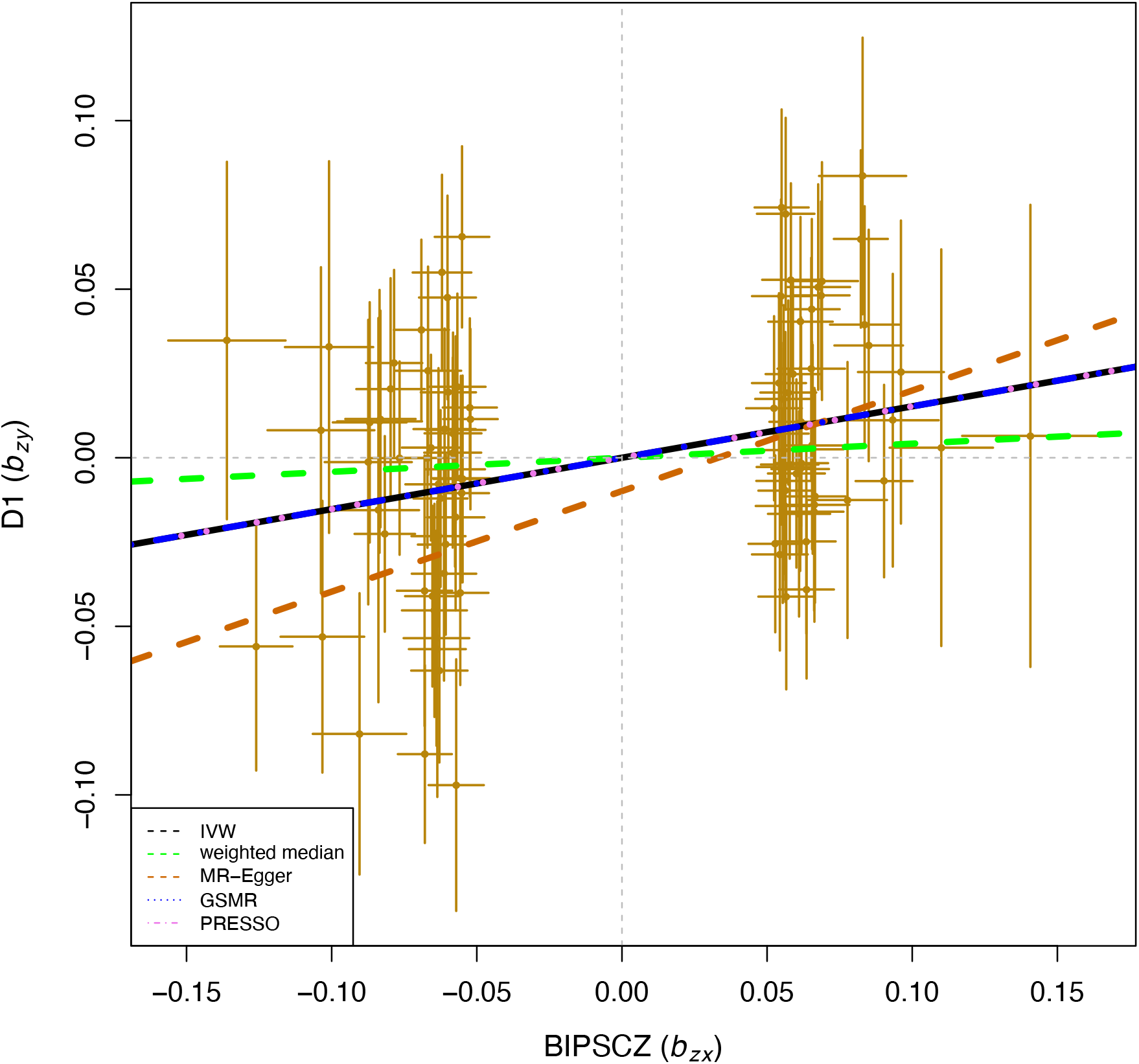
Scatter plot of MR analyses using several models to examine causal relationships of BIP-SCZ genetic liability on self-reported COVID-19. The five models applied in the current manuscript are all depicted. Lines in black, green, brown, blue and purple represent results for fixed-effect IVW, weighted median, MR Egger, GSMR and MR-PRESSO models using 96 instruments. Neither GSMR nor MR-PRESSO identified any instrument outliers. Hence, the MR-PRESSO result was same as the IVW result, which was almost the same as the GSMR result, resulting in overlapping lines in the graph. Error bars represent effect size standard errors.

**Table 3.** Forward MR results of BIP-SCZ as a risk factor for self-reported COVID-19 (D1), using 96 instrument variables.

| MR model | OR | Lower<br>95%CI | Upper<br>95%CI | P-value |
| --- | --- | --- | --- | --- |
| GSMR | 1.165 | 1.062 | 1.277 | 0.0012 |
| Inverse Variance Weighted | 1.162 | 1.052 | 1.283 | 0.0031 |
| MR-PRESSO | 1.162 | 1.052 | 1.283 | 0.0045 |
| Weighted median | 1.045 | 0.91 | 1.2 | 0.5336 |
| MR Egger | 1.349 | 0.816 | 2.23 | 0.2467 |

**Figure 2A.**
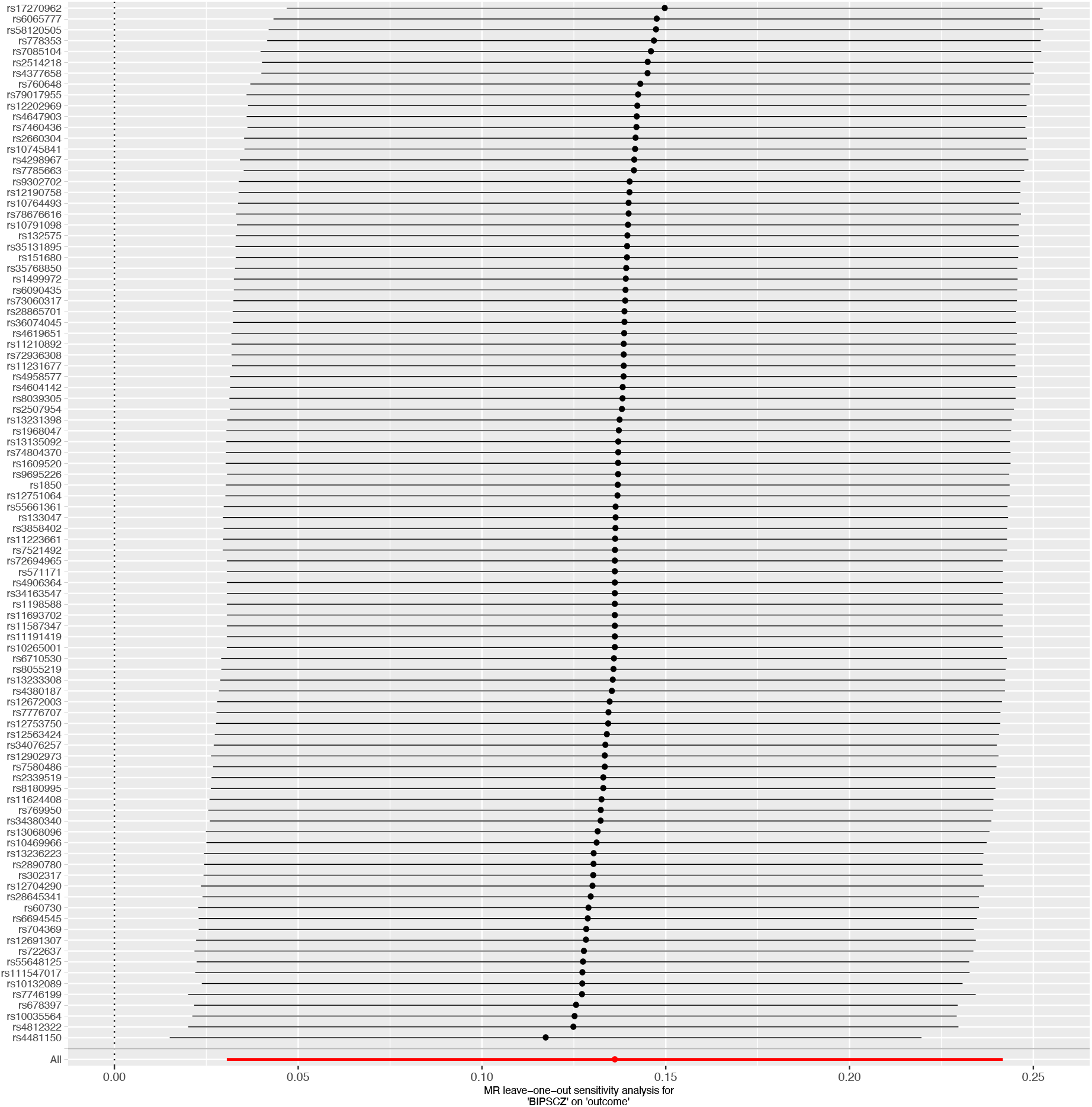
Leave-one-out analysis to evaluate whether any single instrumental variable was driving the causal association of BIP-SCZ with self-reported COVID-19 disproportionately. As can be appreciated from the graph, no genetic variant altered the pooled beta coefficient, indicating stability of our results.

**Figure 2B.**
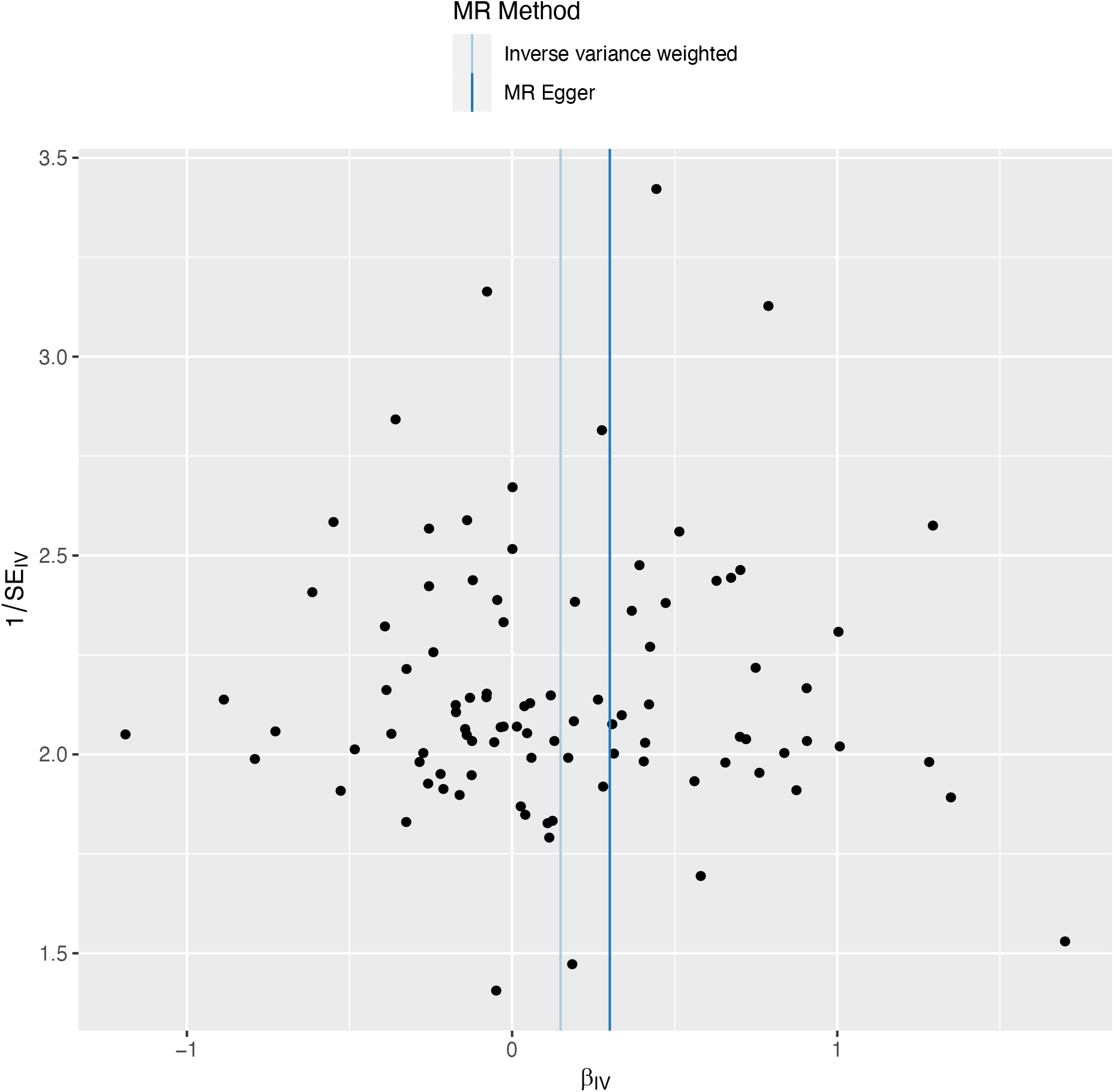
Generalized funnel plot of univariable MR analysis of BIP-SCZ genetic liability effects on self-reported COVID-19 with first-order IVW and MR-Egger regression slopes to look for asymmetry as a sign of pleiotropy. This kind of graph plots the ratio estimate for each variant on the horizontal axis against its square-root precision (or weight) on the vertical axis. As can be appreciated from the plot, no evidence for asymmetry was detected, indicating absence of directional pleiotropy and the instrument strength independent of direct effect (InSIDE) assumption to be satisfied.

At a nominally significant p-value threshold (p<0.05), we detected causal effects of genetic liability to SCZ on very severe COVID (A2) and COVID-19 susceptibility (D1), of genetic liability to BIP on severe and very severe COVID-19 (A1 and B2), and of genetic liability to AD on COVID-19 susceptibility (C1 and D1, **Supplementary table 3A**). In line with the significant GSMR finding for BIP-SCZ, all of these nominally significant results had positive odds ratios for (neuro)psychiatric disorders on COVID-19 phenotypes (**Supplementary table 3A;** see **Supplementary Table 3B** for sensitivity analysis and **Supplementary Table 4** for reverse results). The other 4 MR models’ results were consistent with the GSMR findings (**Supplementary Table 5**). Sensitivity analyses using p-value <10^−7^ to select more genetic instruments yielded consistent results, i.e. only genetic liability to BIP-SCZ significantly increased risk to COVID-19, while liabilities to SCZ, BIP and AD increased risk at nominally significant levels (**Supplementary table 3B**).

### Reverse MR results of COVID-19 phenotypes as potential risk factors of neuropsychiatric disorders

Three COVID-19 phenotypes met our predefined cut-off for inclusion as exposures, i.e. GWASs with ≥2 genome-wide significant hits, in univariable MR analysis, namely severe and very severe COVID-19 (A2, B2) as well as COVID-19 susceptibility (C2, **Table 2**). MDD did not have enough overlapping SNPs to extract ≥2 instrument variables, resulting in 18 tests performed in reverse analyses (6 for A2, B2 and C2; **Table 2**; and **Supplementary Table 4A**). The p-value cut-off for significance was thus Bonferroni corrected to 0.05/18 < 0.0028. We found no results withstanding this multiple testing correction. Among most significant results were: genetic liability to very severe COVID-19 (A2) increasing risk of BIP-SCZ (GSMR OR, 1.036, 95% CI, 1.002-1.071; p = 0.04) and of SCZ (GSMR OR, 1.037, 95% CI, 1.001-1.073; p = 0.042; **Supplementary Table 4A**). The other 4 MR models’ results were in line with the GSMR results (**Supplementary Table 5)**. Sensitivity analyses using p-value <10^−7^ to select more genetic instruments also yielded consistent results (**Supplementary table 4B**).

### Multivariable MR analyses

In forward MVMR, we examined the potential effects of genetic liabilities to three (neuro)psychiatric phenotypes jointly (AD, BIP, and SCZ; see methods) on seven COVID-19 phenotypes, resulting in a Bonferroni corrected (for the number of exposures) significance threshold of p < 0.05 / 7 = 0.0071 (**Supplementary Table 6A**). No (neuro)psychiatric disorder showed a robust relationship with COVID-19 in MV-MR-IVW models. Genetic liability to AD, BIP and SCZ showed a nominally significant causal relationship with COVID-19. However, the estimates were not consistent with estimates from the MVMR-Egger sensitivity analysis, where no p-values were < 0.05 (**Supplementary Table 6A**).

For reverse multivariable MR analyses examining the potential effects of genetic liabilities to three COVID-19 phenotypes jointly (A2, B2 and C2, i.e. GWASs with ≥2 genome-wide significant hits) on (neuro)psychiatric phenotypes, no COVID-19 phenotypes showed a causal relationship with any of neuropsychiatric disorders at a Bonferroni-corrected or nominal significance level in both IVW and MR-Egger (**Supplementary Table 6B**). The estimates were consistent with estimates from the MVMR-Egger sensitivity analyses (**Supplementary Table 6B**).

In sensitivity analysis #2 using univariable GSMR analyses we found BMI, EA, CP ad CAD had causal effects on COVID-19 sub phenotypes (**Supplementary Table 7**). When adding all conditions (namely BMI, EA, CP, MDD, BIPSCZ, CAD, CDG, BIP, SCZ, AD and T2D) into MV-MR models, causal effects were observed only between BMI and COVID-19 phenotypes (A2, B2 and C2). MV-MR-IVW model results were consistent with MV-MR-Egger, meaning no neuropsychiatric disorder showed a robust relationship with COVID-19 in these MV-MR models (**Supplementary Table 8**).

In the end, we estimated genetic heritability and genetic correlations between (neuro)psychiatric phenotypes, COVID-19 (Table 1) and BMI. We show these in **Supplementary Table 9**. For (neuro)psychiatric phenotypes we found only nominally significant (p<0.05) genetic correlations. At a Bonferroni-corrected significance threshold (0.05/61=0.00082) we found only significant correlations between BMI and COVID-19 phenotypes. In addition, the LDSC intercepts were close to 0, further confirming absence of substantial sample overlap between GWASs in this MR study.

## Discussion

We evaluated potential bidirectional associations between (neuro)psychiatric diseases and COVID-19 susceptibility and severity. While our univariable GSMR results hint that genetic liability to a combined phenotype of bipolar disorder and schizophrenia could slightly increase susceptibility to COVID-19, other methods including MVMR did not confirm this finding.

Our findings of no consistently increased risks of genetic liabilities to psychiatric disorders on COVID-19 are consistent with recent epidemiological observations in the UK and South Korea ^1,4^ but inconsistent with other reports ^2,3,5^. Similarly, contrary to one study ^2^, we found no evidence of COVID-19 influencing risk of psychiatric disorders. Also in contrast to previous studies, we found no evidence of certain psychiatric disorders increasing risk to develop COVID-19, e.g. of anxiety disorders ^3,5^. As pointed out by the authors of such studies, lack of repeated measures may have resulted in misclassification of important covariates resulting in invalid correction for some covariates in one or more of those studies. Moreover, residual confounding due to some unmeasured variables such as population stratification may result in overestimations of effect sizes in observational studies.

Furthermore, BMI was either not included at all or not included as a continuous covariate in studies reporting positive associations between psychiatric disorders and COVID-19^2,4^. As BMI may be the strongest risk factor for COVID-19 susceptibility (as also confirmed here), it is of interest to correct for BMI as a continuous trait in upcoming epidemiological studies of COVID-19 and psychiatric disorders. Finally, our findings of mostly absent associations or in the event of suggestive associations odds ratios slightly above 1 are in disagreement with some observational studies but in line with MR studies of other COVID-19 risk factors^6-8,41^. Strengths of our study include the integration of univariable and multivariable, bidirectional MR analyses using many instrument variables drawn from large GWASs. We also included a range of (neuro)psychiatric as well as COVID-19 phenotypes and validated our results across available MR methods. Furthermore, verification of primary GSMR findings with other MR methods helped elucidate consistency of findings. Finally, employing a multivariable in addition to a univariable approach is a strength. However, our results should also be interpreted in light of several limitations. First, a general concern in MR studies is risk of sample overlap. We minimized chances of sample overlap between exposure datasets and outcome datasets by excluding UKBB populations from (neuro)psychiatric GWASs and by excluding 23andme cohorts from COVID-19 datasets. We confirmed the lack of substantial sample overlap with LDSC intercepts. Nonetheless, cryptic relatedness and potential sample overlap between exposure and outcome GWASs may result in some degree of instrument bias. However, the F-statistics we found were all above 36, allaying major concerns about weak instrument bias. Another limitation directly follows from the availability of GWAS data. For some phenotypes, such as obsessive-compulsive disorder and anorexia nervosa, no large datasets excluding the UKBB were available at the time of analysis or writing. For MDD and SCZ, summary statistics of larger GWASs may become available in the coming year. Similarly, as COVID-19 GWAS sample sizes ramp up, statistical power in MR analysis may increase. We encourage researchers to repeat MR analyses on other phenotypes and to use such larger GWAS datasets once they become available. To that end, we have uploaded our code to Github (see data availability section above). Possibly, increased statistical power in GWASs of psychiatric phenotypes and COVID-19 will in future elucidate more genetic associations and thus empower future MR analyses. On a similar note, non-whites are underrepresented in GWASs. As individuals of Asian and African ancestries seem particularly prone to develop COVID-19 ^42^, we hope future GWAS encompassing multi-ethnic cohorts will allow for more inclusive MR analyses. Finally, inherent in a design where univariable as well as multivariable, bidirectional analyses are performed, a large number of statistical tests is performed, increasing the multiple-testing burden and statistical penalty.

In conclusion, while genetic liability to bipolar disorder and schizophrenia combined slightly increased COVID-19 susceptibility in univariable GSMR analysis, other MR methods and multivariable analyses could only confirm genetic underpinnings of BMI to be causally implicated in COVID-19 susceptibility. Thus, using MR we found no consistent proof of genetic liabilities to (neuro)psychiatric disorders contributing to COVID-19 liability or vice versa, which is in line with at least two observational studies. As BMI is strongly associated with psychiatric disorders as well as COVID-19, previously reported positive associations between psychiatric disorders and COVID-19 may have resulted from statistical models incompletely capturing BMI as a continuous covariate.

## Data Availability

Data and code availability
We have made our code publicly available on Github (https://github.com/Bochao1/MR_PSY_COVID19). The datasets we accessed to perform our analyses may be found in the publications that listed as references for the datasets used.

## Legends to Supplementary Tables

Supplementary Table 1. The cohort components of the COVID-19 GWASs.

Supplementary Table 2. Additional conditions that were added as exposures for sensitivity analyses given their possible roles in both psychiatric disorders and COVID-19.

Supplementary Table 3 A. Univariable forward GSMR results with nominal significant level (P<0.05) of (neuro)psychiatric disorders on COVID-19 in bold, ordered by decreasing significance.

Supplementary Table 3B. Univariable forward GSMR results using a more lenient p-value threshold (10^−7^) for inclusion of genetic variants of (neuro)psychiatric disorders on COVID-19, with nominal significant level (P<0.05) ordered by decreasing significance.

Supplementary Table 4 A. Univariable reverse GSMR results of (neuro)psychiatric disorders on COVID-19, ordered by decreasing significance.

Supplementary Table 4B. Univariable reverse GSMR results using a more lenient p-value threshold (10^−7^) for inclusion of genetic variants of COVID-19 on (neuro)psychiatric disorders, ordered by decreasing significance.

Supplementary Table 5. Univariable MR results of (neuro)psychiatric disorders and COVID-19 using four other MR models (analyses with p<0.05 in GSMR are depicted, for forward and reverse MR).

Supplementary Table 6A. Forward multivariable MV-MR results of (neuro)psychiatric disorders on COVID-19.

Supplementary Table 6B. Reverse multivariable MV-MR results of COVID-19 on (neuro)psychiatric disorders.

Supplementary Table 7. Univariable forward GSMR results of additional conditions (see Suppl. Table 2) on COVID-19.

Supplementary Table 8. MV-MR results of additional conditions and (neuro)psychiatric disorders on COVID-19.

Supplementary Table 9. SNP-based heritabilities of COVID-19 phenotypes and (neuro)psychiatric disorders & genetic correlations. And genetic correlations within COVID-19 phenotypes and with (neuro)psychiatric disorders.

