## supplements for "Are psychiatric disorders risk factors for COVID-19 susceptibility and severity? a two-sample, bidirectional, univariable and multivariable Mendelian Randomization study"

**Supplementary Figure 1.** Representation of the principle of bidirectional Mendelian Randomization (MR): assessing causal associations of genetic variants that act as genetic instrumental variables, i.e. as proxies for (neuro)psychiatric disorders, with COVID-19 outcomes.
Genetic instruments are genetic variants used as proxies for (modifiable) exposures: (neuro)psychiatric disorders in forward MR analyses and COVID-19-related phenotypes in reverse analyses. The valid genetic variant for IV analysis must be: (1) truly associated with the exposures, so that it can be used as a proxy for exposure of interest; (2) not associated with measured/unmeasured confounders of the exposure-outcome relation; and (3) meet exclusion restriction criteria, i.e., the instrument/genetic variant should not be directly associated with the outcome and its effect must be mediated only through the exposure.
As can be appreciated from panels A and B, genetic instruments allow for assessments of direct associations between genetic instruments as the exposure with an outcome in single (univariable) MR, which is illustrated with the dashed line (e.g. in A: effects of a (neuro)psychiatric disorder on a COVID-19 outcome). By contrast, in observational studies not making use of genetic instruments confounders may (partially) explain the association between two or more traits.
As can be appreciated in panels C and D, Multivariable MR (MVMR) estimates the effects of each exposure on an outcome when multiple phenotypes may be associated with the exposure. Scenarios one may think of are when multiple exposures may be related to one another or when one exposure may mediate the relationship between the exposure of interest and an outcome. MVMR does so by using genetic instruments associated with each of those multiple phenotypes.
A=Forward single MR; B=Reverse single MR; C=Forward multivariable MR (MVMR); D=Reverse MVMR. LD Pruning = Linkage Disequilibrium pruning used to obtain independent genetic instruments. SCZ=schizophrenia; BIP=bipolar disorder; AD=Alzheimer's disease.

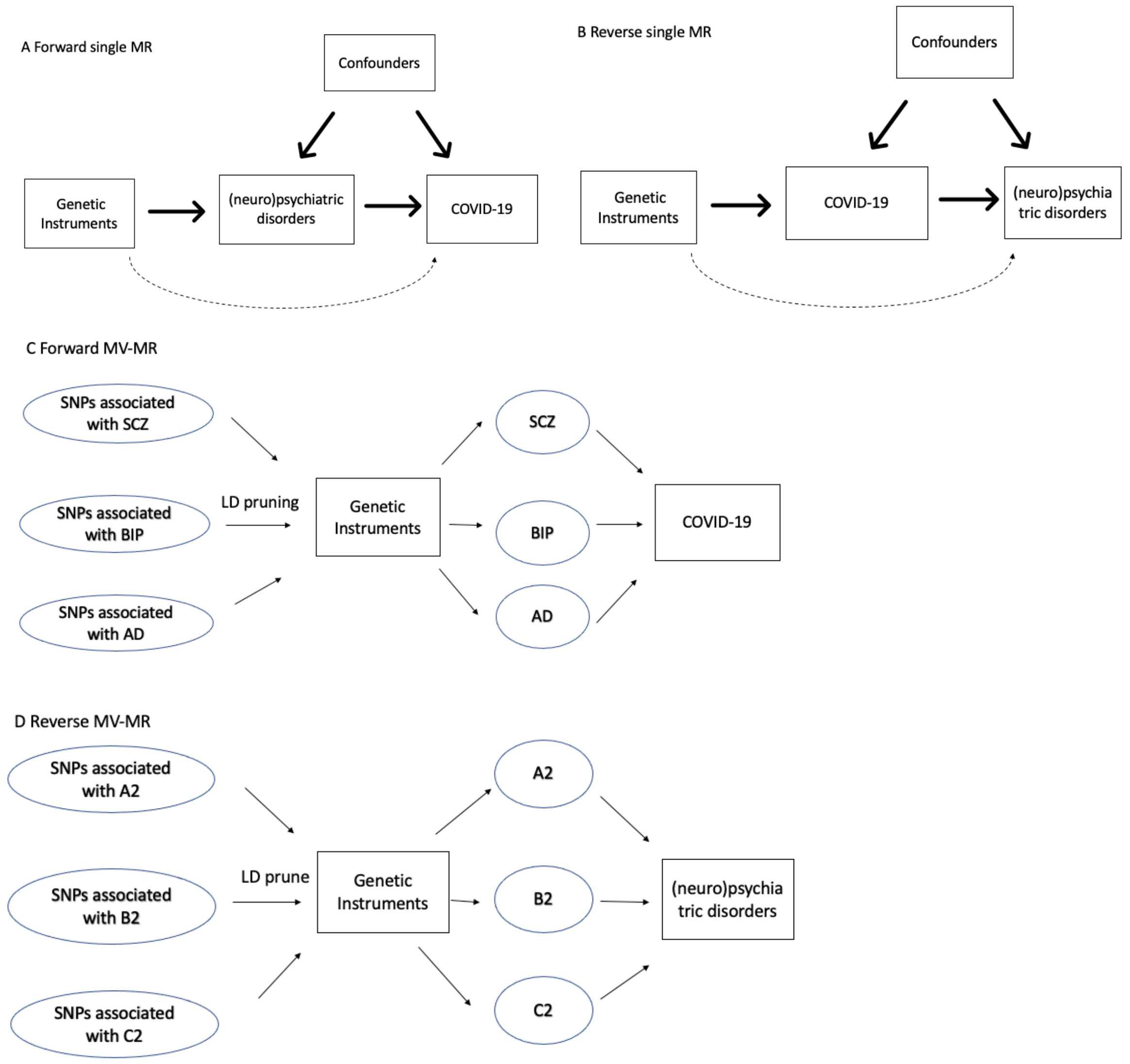

**Supplementary Figure 2.** Scatter plot for the sharing model (left) illustrating the pattern induced by a shared factor (correlated pleiotropy) without a causal effect; scatter plot for the causal model (middle) illustrating the pattern induced by a causal effect; and scatter plot of expected log pointwise posterior density (ΔEPLD), which measures how well the posterior distributions estimated under a given model are expected to predict a hypothetical new set of summary statistics obtained from GWAS BIPSCZ (a combined GWAS of bipolar disorder and schizophrenia) and COVID-19 in different samples.

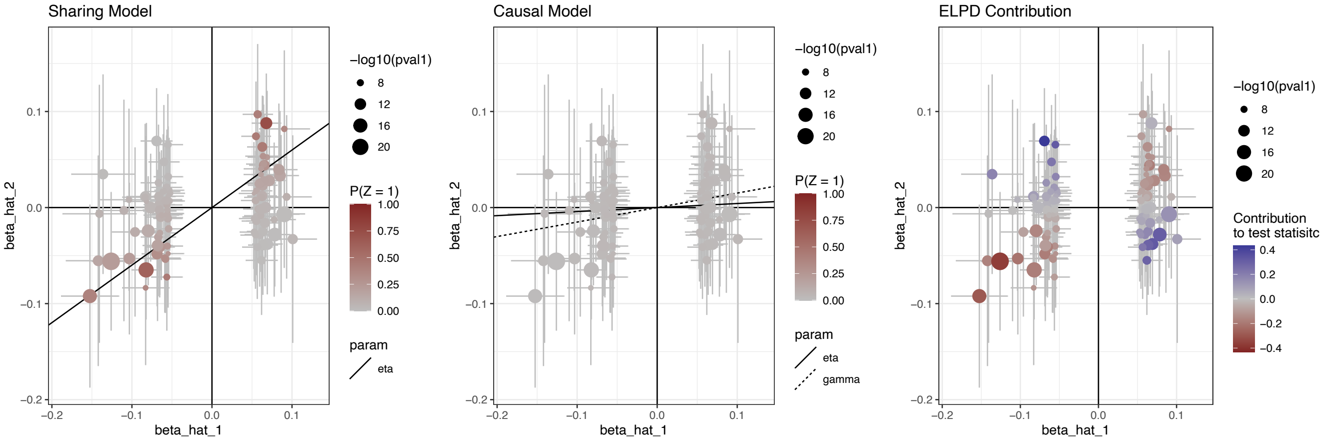


Effect estimates for trait BIPSCZ (horizontal axis) are plotted against estimates for COVID-19 D1 (i.e. predicted COVID-19 diagnosis, vertical axis). Point size is proportional to the p-value for BIPSCZ. Error bars have a length 1.96 times the standard error of the estimate.

**Legends to Supplementary Tables**

Supplementary Table 1. The cohort components of the COVID-19 GWASs.

Supplementary Table 2. Additional conditions that were added as exposures for sensitivity analyses given their possible roles in both psychiatric disorders and COVID-19.

Supplementary Table 3 A. Univariable forward GSMR results with nominal significant level ($P<0.05$) of (neuro)psychiatric disorders on COVID-19 in bold, ordered by decreasing significance.

Supplementary Table 3B. Univariable forward GSMR results using a more lenient p-value threshold ($10^{-7}$) for inclusion of genetic variants of (neuro)psychiatric disorders on COVID-19, with nominal significant level ($P<0.05$) ordered by decreasing significance.

Supplementary Table 4 A. Univariable reverse GSMR results of (neuro)psychiatric disorders on COVID-19, ordered by decreasing significance.

Supplementary Table 4B. Univariable reverse GSMR results using a more lenient p-value threshold ($10^{-7}$) for inclusion of genetic variants of COVID-19 on (neuro)psychiatric disorders, ordered by decreasing significance.

Supplementary Table 5. Univariable MR results of (neuro)psychiatric disorders and COVID-19 using four other MR models (analyses with $p<0.05$ in GSMR are depicted, for forward and reverse MR).

Supplementary Table 6A. Forward multivariable MV-MR results of (neuro)psychiatric disorders on COVID-19.

Supplementary Table 6B. Reverse multivariable MV-MR results of COVID-19 on (neuro)psychiatric disorders.

Supplementary Table 7. Univariable forward GSMR results of additional conditions (see Suppl. Table 2) on COVID-19.

Supplementary Table 8. MV-MR results of additional conditions and (neuro)psychiatric disorders on COVID-19.

Supplementary Table 9. SNP-based heritabilities of COVID-19 phenotypes and (neuro)psychiatric disorders & genetic correlations. And genetic correlations within COVID-19 phenotypes and with (neuro)psychiatric disorders.
